# High Rate of Transplantation Prior to Review of Status Exception Requests among Adult Heart Transplant Candidates

**DOI:** 10.1101/2025.09.12.25335606

**Authors:** Daniel J Ahn, Toshihiro Nakayama, Antony Attia, Molly White, Dalin Eap, Nikhil Narang, Kiran K Khush, William Parker, Kazunari Sasaki

## Abstract

**Background:** In the United States heart allocation system, when transplant centers submit applications for status exceptions to increase waitlist priority, patients obtain the requested status upgrades immediately while their applications are sent to the regional review boards (RRBs) and reviewed retrospectively. How much time elapses between obtaining a status upgrade through exception and application receipt by the RRBs and how often transplants occur during this period is unknown.

**Methods:** Using the Scientific Registry of Transplant Recipients (SRTR), we identified all adult heart transplant candidates listed between October 18, 2018 and December 31, 2023 with submitted applications for status exceptions. We assessed 1) the amount of time elapsed between submission of exception applications and their receipt by the RRBs and 2) the rate of heart transplantation during this “travel” time, stratified by whether the applications were eventually approved or denied. Additionally, using complete match run data, we estimated how many listed patients were skipped by candidates who received transplants with exceptions that were ultimately denied.

**Results:** 135 transplant centers submitted status exception requests on behalf of 8,269 adult candidates during the study period, of whom 608 (7.4%) received a denial at least once. The median time from obtaining higher priority statuses immediately via exceptions to application receipt by the RRBs was 3 days. 2,087 out of 8,269 (25.2%) patients received transplants before the RRBs even received their applications, with 115 (18.9%) among 608 with eventual denials and 1,972 (25.7%) among 7,661 with approvals. The cumulative incidence of heart transplantation before application receipt for eventual denials was 19.1% (95% CI [16.0%, 22.3%]) and that for approvals was 26.2% (95% CI [25.2%, 27.1%]) (p < 0.001) at 2 weeks. Based on match run data, the 115 patients who received transplants with denied exceptions bypassed more than seven thousand potential transplant recipients.

**Conclusions:** More than 25% of patients with status exception requests receive heart transplants before their applications are even received by their respective RRBs, let alone reviewed. This raises significant concerns about the efficacy and fairness of retrospective review of exception requests for the allocation of valuable donor hearts.

## Introduction

Since October 2018, the United States (US) donor heart allocation system has rank-ordered adult transplant candidates using six ordinal statuses. To qualify for the highest priority statuses, most patients must have objective hemodynamic or laboratory evidence of cardiogenic shock.^1^ However, the system allows transplant centers to request exceptions on behalf of select candidates. In exception applications, centers may argue to their respective regional review board (RRB) that their patients have a similar level of medical urgency as others who have qualified for the same status by meeting standard criteria. One of the goals of the 2018 heart allocation policy change was to reduce the system’s dependence on exception listings by increasing the number of available statuses.^2^ However, the proportion of listings with exceptions has increased dramatically to about 30-40%.^3–7^ Additionally, the approval rate for heart exception requests is consistently greater than 95%, with most denials eventually getting approved through appeals.^3^ Worryingly, candidates listed with status exceptions have significantly reduced waitlist mortality compared to those who are listed via standard criteria.^8,9^

As the heart transplant community seeks to limit the overuse of exceptions, one aspect of exception allocation that has not been previously studied is how exception requests are reviewed retrospectively. Upon submission of exception requests, all applicants immediately receive the requested status and accrue high-priority listing time while the requests are still in transit to the RRBs and are either approved or denied retrospectively through a voting process that may take up to a week.^10^ If denied, transplant centers may submit appeals to the RRB, and if still unfavorable, to the Organ Procurement and Transplantation Network (OPTN) Heart Committee, all the while patients continuing to possess the requested status.^1^ In this study, we determined how much time candidates spend with status upgrades before the RRBs receive their exception applications and how often heart transplants occur during this period.

## Methods

### Data Source and Study Population

This study used data from the Scientific Registry of Transplant Recipients (SRTR). The SRTR includes data on all donors, waitlisted patients, and transplant recipients in the United States, submitted by the members of the OPTN. The Health Resources and Services Administration (HRSA) provides oversight to the activities of the OPTN and SRTR contractors. This study was exempted by the University of Chicago and Stanford University Institutional Review Boards.

We identified all adult heart transplant patients in the SRTR dataset listed between October 18, 2018 and December 31, 2023 who received a status exception at least once, with follow-up until March 31, 2024. We collected relevant demographic and clinical data from patients at initial listing. Variables included sex, age at listing, body mass index (BMI), race, blood type, primary diagnosis, and mechanical circulatory support devices. We categorized BMI into the following: underweight (<18.5 kg/m^2^), normal (18.5-24.9 kg/m^2^), overweight (25-29.9 kg/m^2^), and obese (≥30 kg/m^2^).

### Outcomes and Analysis

The primary outcome of the study was heart transplantation. The secondary outcome was the time elapsed between patients obtaining status exceptions and the RRBs receiving the respective exception applications for review. Because the date of final approval or denial by the RRB is not included in the files provided by the SRTR, we were unable to determine the time elapsed between exception application submission and final review by the RRB. A graphical representation of the exception review process is in **Figure 1**.

**Figure 1:**
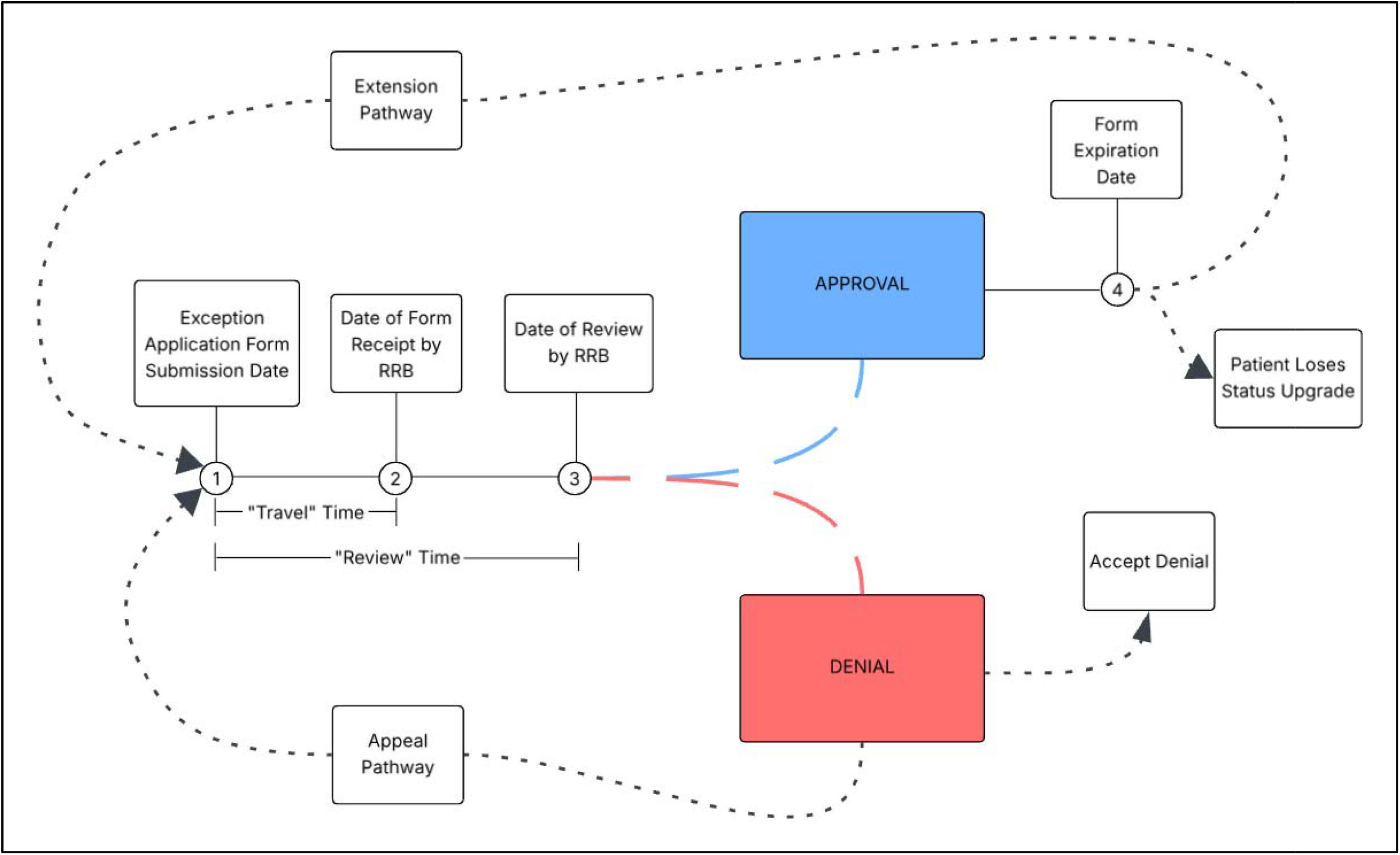
Graphical representation of the retrospective review process of applications for status exceptions in the adult heart allocation system. Listing centers may submit initial applications for status exceptions at Timepoint 1, and patients obtain the requested statuses immediately. After a “travel” time, the regional review boards (RRBs) receive the applications at Timepoint 2 and make a final decision through a majority voting process at Timepoint 3, which completes the “review” time. If approved, patients keep the requested status they obtained at Timepoint 1 until their status upgrade expires at Timepoint 4, which is based on the status (e.g. 7 days for status 1). Before Timepoint 4, centers may either allow the approved forms to expire and have patients lose the upgraded status or submit applications to extend the exceptions, which restarts the review process at Timepoint 1. If initial applications are denied at Timepoint 3, centers may accept the denial, and patients drop to the status they meet through standard criteria. Alternatively, centers may submit forms for appeal within a required amount of time, which causes the review process to start again at Timepoint 1. Because review of exception applications is retrospective, patients always have the requested status throughout the “review” time, even if their applications are denied at Timepoint 3. Furthermore, while undergoing the “appeal” pathway, candidates possess the same requested status they had since submission at Timepoint 1 even with denial by the RRBs.

We first identified all initial exception applications (for initial listings and status extensions) that were submitted and then calculated the approval rate, stratified by UNOS region. We also calculated the proportion of all waitlisted candidates who submitted exception applications, stratified by transplant center. Next, we calculated the amount of time that elapsed between patients obtaining status designations through exceptions and the receipt of the exception applications by the respective RRBs. We stratified this “travel” time by both requested status on the waitlist and UNOS region. We compared the median “travel” time by region for each status with the Kruskal-Wallis test.

Furthermore, we identified all heart transplant candidates who received a heart transplant while their exception applications were still en route to the RRBs, stratified by whether they were eventually approved or denied. For the patients who received transplants with exceptions that were eventually denied, we estimated what status they would have had at the time of transplant without the exceptions. To perform this analysis, for patients who were already on the waitlist, we simply carried forward the status they had before submitting a status exception. For those who were inactive immediately before obtaining an exception or who received an exception at the time of initial listing, we determined what status they would have had based on OPTN heart policy criteria using information on therapies such as mechanical circulatory support and hemodynamic measurements documented in the status justification files.

We next utilized complete match run data provided by the OPTN to estimate how many potential transplant recipients (PTRs) were skipped due to candidates getting listed at a higher status by exceptions that were eventually denied. For simplicity of analysis, we did not calculate waiting time to determine exact predicted placement in the match run. Instead, we assumed that for given predicted statuses without exceptions, candidates were at the top of their respective sequence classification in the match run. In effect, this provides the most conservative estimate of the true number of “skipped” PTRs. For the “skipped” PTRs, we then estimated the cumulative incidence of death or removal for clinical deterioration within 6 weeks of the match run, treating transplantation as a competing event.

We also calculated the cumulative incidence (CI) of transplant before the RRBs received applications for exception requests within 2 weeks. In this analysis, there were two competing events: 1) death or waitlist removal for clinical deterioration before application receipt and 2) application receipt by the RRBs. We chose 2 weeks as this was the longest recorded time from submission to receipt in the dataset. We then compared the respective rates, stratified by exception approval or denial, with Fine-Gray analysis. Because the SRTR lacks data on date of review, we performed a sensitivity analysis estimating the CI of transplant by 3 days after application receipt. We chose 3 days because after an RRB receives an application, the primary representative assigned to the case must vote within 3 days, after which all RRB members cast votes.^10^ We then calculated how many more heart transplants occurred with this change. Finally, to compare demographic and clinical data of patients by approval or denial of exceptions, we performed descriptive statistics with chi-square tests for categorical variables and Wilcoxon rank-sum tests for continuous variables. All statistical tests were two-sided, and we considered a p-value of < 0.05 to be significant. We performed all analyses with R (version 4.5.1). Complete statistical code necessary to reproduce the study results is available online.^11^

## Results

Between October 18, 2018 and December 31, 2023, 38,594 adult patients were added to the heart transplant waitlist at 142 US transplant centers, of whom 8,269 patients at 135 centers had at least one status exception request submitted on their behalf. The per-center rate of candidates with submitted exceptions differed significantly by transplant center from 0% to 100%, p < 0.001 (median 25.2%) (**Supplemental Figure 1**). 7,661 patients (92.6%) had all exception applications approved by their RRBs, while 608 (7.4%) submitted at least one exception application that was denied by the RRBs (**Table 1**). Patients with denials were more likely to be male (77.5% vs 72.5%, p = 0.008), have blood type O (56.3% vs 46.7%, p < 0.001), have requested higher statuses (status 1: 15.8% vs 12.8%, status 2: 61.3% vs 58.3%, p < 0.001), and have lower ECMO (4.5% vs 21.3%, p < 0.001) and higher LVAD (53.2% vs 39.1%, p < 0.001) utilization at listing. There were no significant differences in race or primary diagnosis. The most common diagnosis was non-ischemic dilated cardiomyopathy (37.9%).

**Table 1:** Demographic and Clinical Characteristics of Study Population, Stratified by Approval and Denial of Exception Applications

| Characteristic | Overall<br>N = 8,269<sup>1</sup> | Approved<br>N = 7,661<sup>1</sup> | Not Approved<br>N = 608<sup>1</sup> | p-value<sup>2</sup> |
| --- | --- | --- | --- | --- |
| Age at Listing (Years) | 56 (44, 63) | 56 (44, 63) | 56 (45, 62) | 0.7 |
| Sex |  |  |  | 0.008 |
| Male | 6,027 (72.9%) | 5,556 (72.5%) | 471 (77.5%) |  |
| Female | 2,242 (27.1%) | 2,105 (27.5%) | 137 (22.5%) |  |
| Race |  |  |  | 0.092 |
| White | 4,655 (56.3%) | 4,321 (56.4%) | 334 (54.9%) |  |
| Black | 2,319 (28.0%) | 2,160 (28.2%) | 159 (26.2%) |  |
| Hispanic/Latino | 816 (9.9%) | 741 (9.7%) | 75 (12.3%) |  |
| Asian | 311 (3.8%) | 281 (3.7%) | 30 (4.9%) |  |
| Other | 168 (2.0%) | 158 (2.1%) | 10 (1.6%) |  |
| BMI |  |  |  | 0.3 |
| Normal (18.5 - 24.9 kg/m<sup>2</sup>) | 2,217 (26.8%) | 2,068 (27.0%) | 149 (24.5%) |  |
| Underweight (< 18.5 kg/m<sup>2</sup>) | 98 (1.2%) | 93 (1.2%) | 5 (0.8%) |  |
| Overweight (25 - 29.9 kg/m<sup>2</sup>) | 2,837 (34.3%) | 2,627 (34.3%) | 210 (34.5%) |  |
| Obese (≥ 30 kg/m<sup>2</sup>) | 3,051 (36.9%) | 2,809 (36.7%) | 242 (39.8%) |  |
| Unknown | 66 (0.8%) | 64 (0.8%) | 2 (0.3%) |  |
| Blood Type |  |  |  | <0.001 |
| O | 3,923 (47.4%) | 3,581 (46.7%) | 342 (56.3%) |  |
| A | 2,800 (33.9%) | 2,638 (34.4%) | 162 (26.6%) |  |
| B | 1,208 (14.6%) | 1,124 (14.7%) | 84 (13.8%) |  |
| AB | 338 (4.1%) | 318 (4.2%) | 20 (3.3%) |  |
| Primary Diagnosis |  |  |  | 0.056 |
| Dilated Cardiomyopathy, Non-Ischemic | 3,124 (37.8%) | 2,900 (37.9%) | 224 (36.8%) |  |

| **Characteristic** | **Overall**<br>**N = 8,269<sup>1</sup>** | **Approved**<br>**N = 7,661<sup>1</sup>** | **Not Approved**<br>**N = 608<sup>1</sup>** | **p-value<sup>2</sup>** |
| --- | --- | --- | --- | --- |
| Ischemic Cardiomyopathy | 2,084 (25.2%) | 1,914 (25.0%) | 170 (28.0%) |  |
| Restrictive Cardiomyopathy | 1,407 (17.0%) | 1,300 (17.0%) | 107 (17.6%) |  |
| Hypertrophic Cardiomyopathy | 315 (3.8%) | 289 (3.8%) | 26 (4.3%) |  |
| Congenital Heart Disease | 550 (6.7%) | 527 (6.9%) | 23 (3.8%) |  |
| Other | 789 (9.5%) | 731 (9.5%) | 58 (9.5%) |  |
| **Requested Status** |  |  |  | <0.001 |
| Status 1 | 1,080 (13.1%) | 984 (12.8%) | 96 (15.8%) |  |
| Status 2 | 4,842 (58.6%) | 4,469 (58.3%) | 373 (61.3%) |  |
| Status 3 | 1,234 (14.9%) | 1,131 (14.8%) | 103 (16.9%) |  |
| Status 4 | 1,113 (13.5%) | 1,077 (14.1%) | 36 (5.9%) |  |
| **Mechanical Circulatory**<br>**Support at Initial Listing** |  |  |  | <0.001 |
| ECMO | 277 (20.5%) | 274 (21.3%) | 3 (4.5%) |  |
| TAH | 8 (0.6%) | 7 (0.5%) | 1 (1.5%) |  |
| IABP | 1,039 (76.8%) | 979 (76.1%) | 60 (90.9%) |  |
| LVAD and RVAD | 28 (2.1%) | 26 (2.0%) | 2 (3.0%) |  |
| LVAD Only | 907 (40.0%) | 832 (39.1%) | 75 (53.2%) |  |
| RVAD Only | 8 (0.4%) | 8 (0.4%) | 0 (0.0%) |  |

<sup>1</sup> Median (Q1, Q3); n (%)
<sup>2</sup> Wilcoxon rank sum test; Pearson's Chi-squared test; Fisher's exact test

### Elapsed Time from Use of Exception to Receipt of Exception Application by RRBs

There were 21,596 exception applications submitted (both requests for initial listing and extensions), with a 96.9% approval rate overall. The approval rate varied significantly by UNOS region from 95.2% in region 8 to 97.8% in region 4 (p < 0.001) (**Supplemental Figure 2, Supplemental Figure 3**) and by transplant center from 66.7% to 100% (p < 0.001). The number of applications for statuses 1 to 4 were 1,476 (6.8%), 10,693 (49.5%), 5,905 (27.3%), and 3,522 (16.3%), respectively. The median “travel” time for all status exception applications to be received by the RRBs was 3 days. The “travel” time varied significantly by region for all four status designations (status 1, p = 0.003; statuses 2 to 4, p < 0.001) (**Figure 2**). The combined “travel” time for patients with denied applications was 3,393 days (9.3 years) and 65,510 days (179.4 years) for patients with approvals.

**Figure 2:**
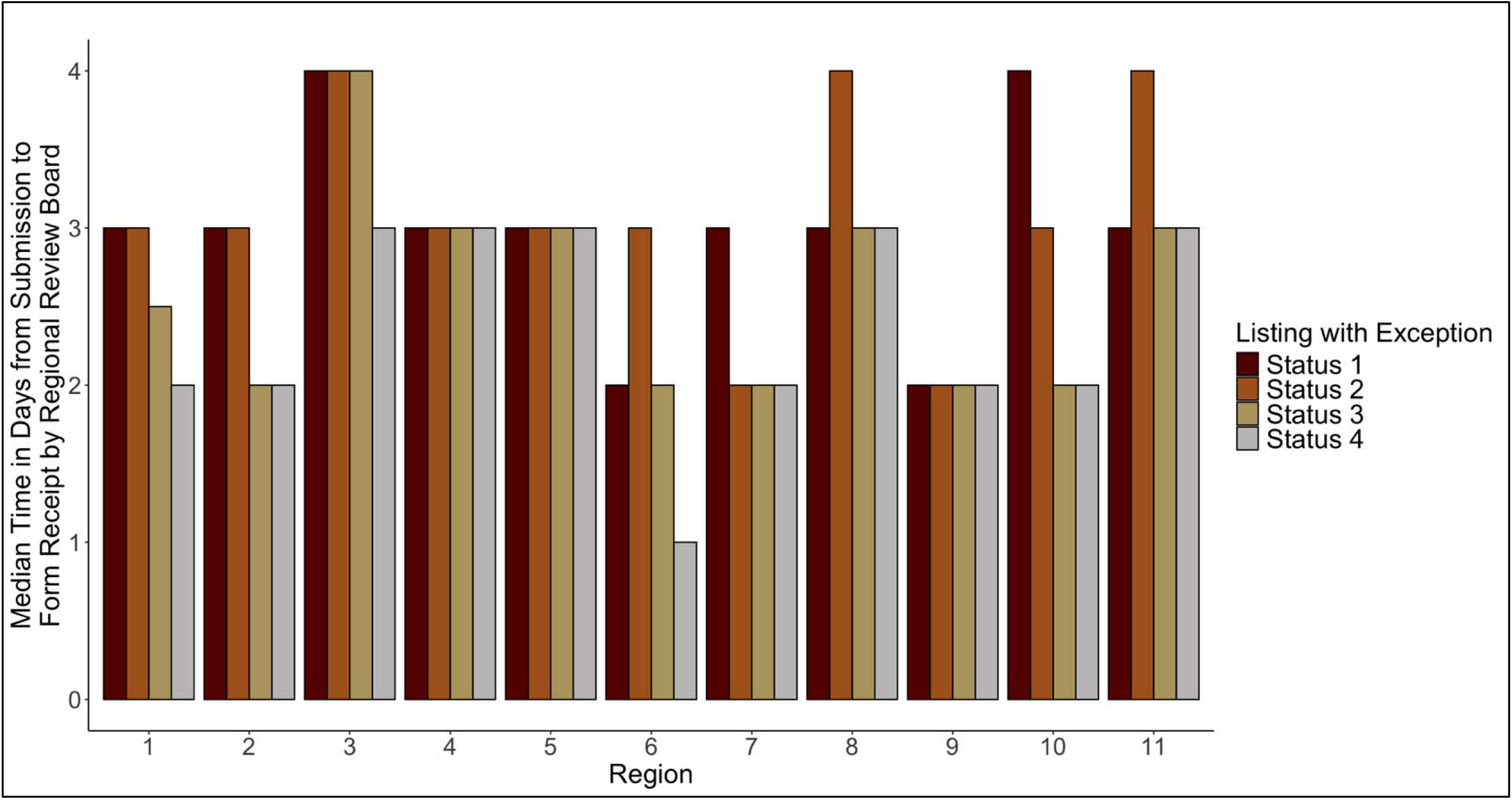
Distribution of median time from exception application submission to application receipt, stratified by UNOS region and requested status upgrade. The median “travel” time overall and by each requested status was 3 days. The “travel” time varied significantly by UNOS region for all 4 status designations (status 1, p = 0.003; statuses 2 to 4, p < 0.001).

### Heart Transplantation before Receipt of Exception Applications by RRBs

For the 608 patients with denied exception applications, the CI of heart transplantation before application receipt was 19.1% (95% CI [16.0%, 22.3%]) at 2 weeks while that for the 7,661 patients with approvals was 26.2% (95% CI [25.2%, 27.1%]) (p < 0.001) (**Figure 3**). The CI of death or removal for deterioration for denied patients was 1.0% (95% CI [0.4%, 2.1%]) and for approved was 1.1% (95% CI [0.9%, 1.4%]) (p = 0.77). 2,087 out of 8,269 (25.2%) patients received transplants before the RRBs even received their applications. During the “travel” time, 115 (18.9%) among 608 patients with eventual denials received a transplant (35 with status 1, 67 with status 2, 12 with status 3, and 1 with status 4), and 1,972 (25.7%) among 7,661 with eventual approvals were transplanted (**Figure 4A**). 91 (1.1%) deaths or removals for deterioration occurred during the “travel” time. The distribution of which statuses the 115 patients should have had at the time of transplant is in **Figure 4B**, stratified by the statuses that they temporarily achieved with their exception requests. 24 (68.6%) out of 35 patients with status 1 would have had status 2. 32 (47.7%) and 19 (28.4%) out of 67 patients transplanted with status 2 exceptions would have had status 4 and 6, respectively.

**Figure 3:**
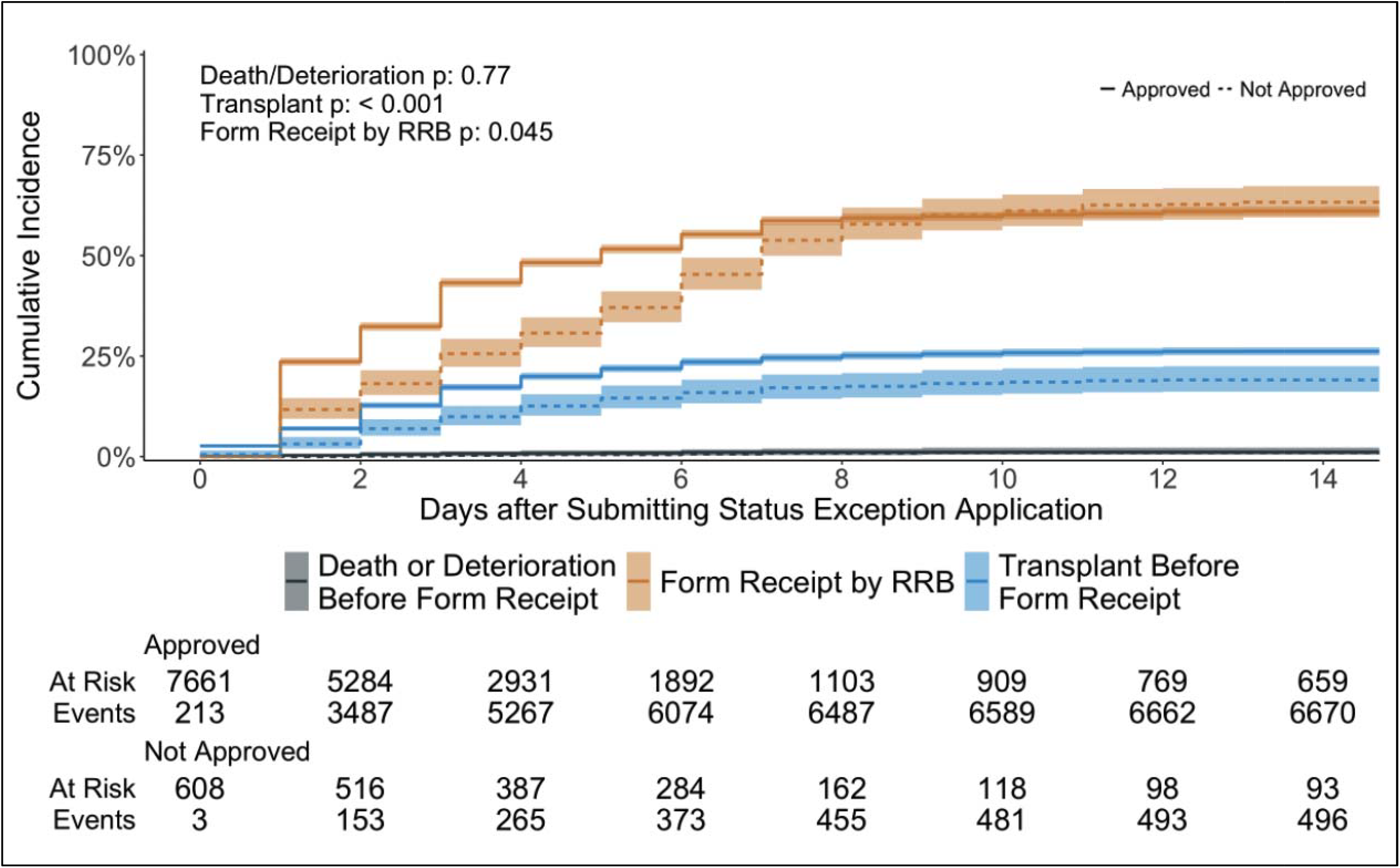
Heart transplantation before receipt of applications for status exceptions by RRBs. This shows the cumulative incidence of transplant before application receipt by the RRBs, treating death or waitlist removal for clinical deterioration before application receipt and receipt of applications by the RRBs as competing events. Plots for application approvals are denoted by solid lines and those for denials by dotted lines. Fine-Gray analysis results for all competing events stratified by approval or denial of exception applications are in the top-left corner.

**Figure 4.**
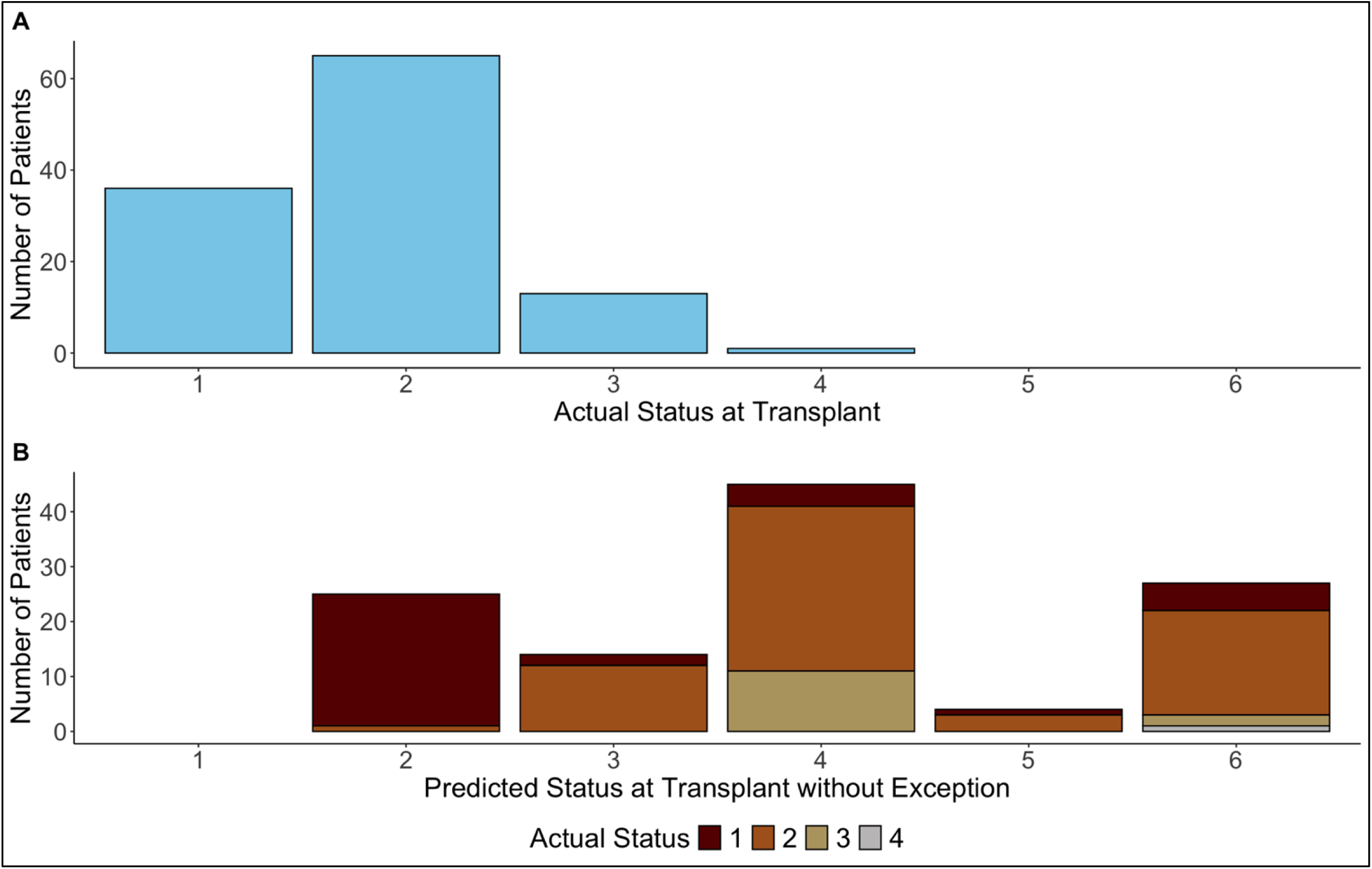
Distribution of actual and predicted statuses without exceptions at transplant. 4A shows the actual distribution of status levels at transplant of the 115 patients who received a transplant with exceptions that were ultimately denied by the RRBs. 4B shows the distribution of the 115 patients’ predicted statuses without exceptions at the time of transplant based on standard criteria described in OPTN policy documents, stratified by what their statuses actually were in 4A.

### Match Run Analysis

If the patients who received prioritized transplants with exceptions that were ultimately denied had remained in their expected status at the time of their match runs, the 115 donor hearts could have been offered to 7,381 other candidates. Within 6 weeks of the match run dates, 117 (1.6%) of the 7,381 “skipped” candidates died or were removed for clinical deterioration, and 2,326 (31.5%) received heart transplants. The cumulative incidence of death or removal for clinical deterioration within 6 weeks of the match runs was 1.6% (95% CI [1.4%, 1.9%]) while that for transplant was 32.0% (95% CI [31.0%, 33.1%]) (p < 0.001) (**Supplemental Figure 4**).

### Sensitivity Analysis

The number of transplants before an estimated review date of 3 days after application receipt for denied and approved patients was 152 (+37, increased by 32.1%) and 3,338 (+1,366, increased by 69.3%), respectively. The total of pre-review deaths and removals for deterioration was 144 (+53, increased by 58.2%). The CI of transplant before estimated RRB review increased for patients with denials at 25.0% (95% CI [21.6%, 28.5%]) and with approvals at 44.2% (95% CI [43.1%, 45.3%]) at 2 weeks (p < 0.001) (**Figure 5**). The CI of death or removal for clinical deterioration before estimated review remained low for patients with denials at 1.2% (95% CI [0.5%, 2.3%]) and for those with approvals at 1.8% (95% CI [1.5%, 2.1%]) at 2 weeks (p = 0.24).

**Figure 5:**
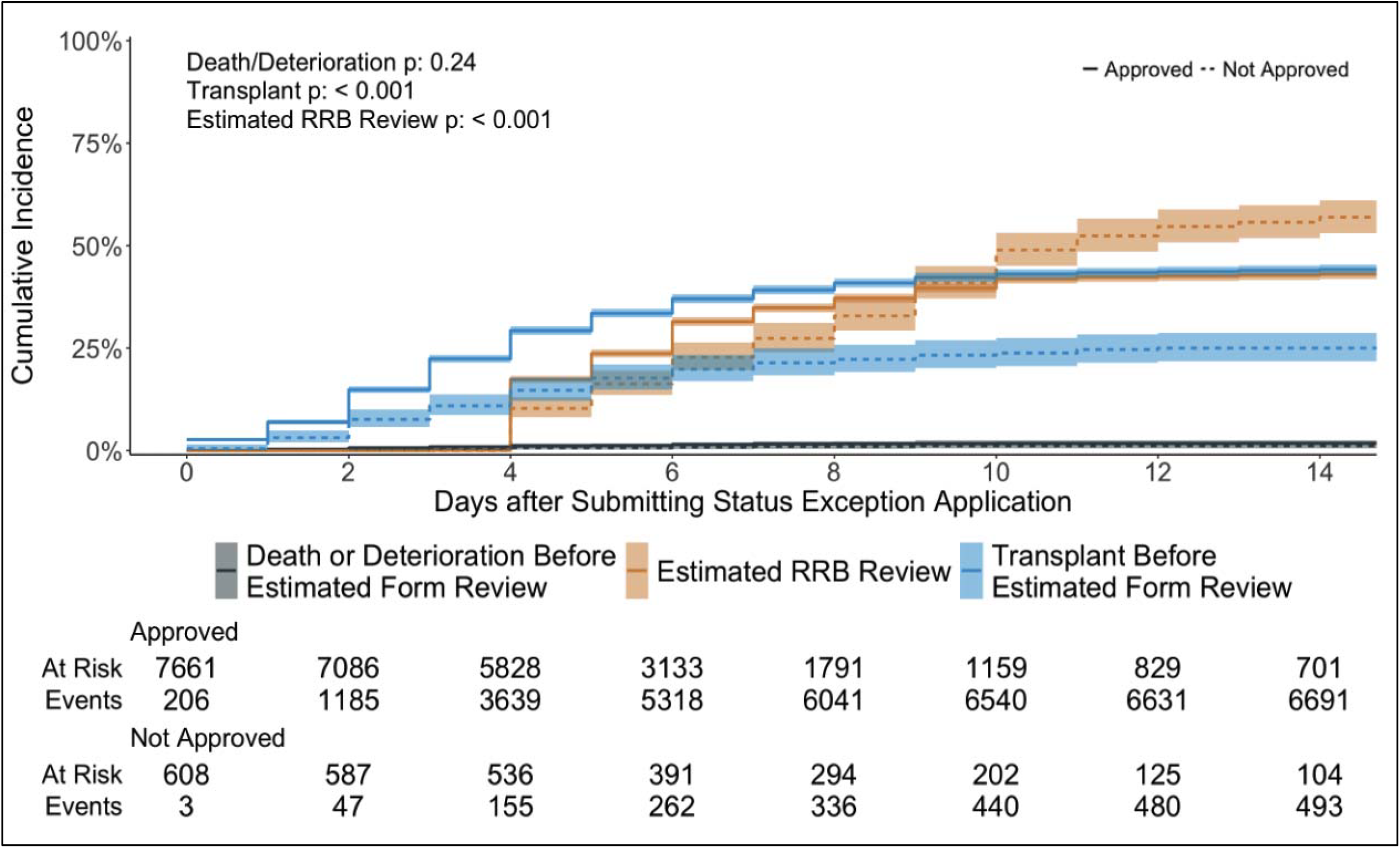
Heart transplantation before estimated review of status exceptions by RRBs. This shows the cumulative incidence of transplant before an estimated review date of 3 days after application receipt by the RRBs, treating death or waitlist removal for clinical deterioration before application receipt and receipt of applications by the RRBs as competing events. Plots for appl cation approvals are denoted by solid lines and those for denials by dotted lines. Fine-Gray analysis results for all competing events stratified by approval or denial of exception applications are in the top-left corner.

## Discussion

In this registry-based study of waitlisted patients awaiting heart transplantation with status exceptions in the US, we report the time elapsed between obtaining exceptions and receipt of applications by the RRBs as well as the rate of heart transplantation while applications are still “in transit” to the RRBs. Our main findings are as follows: 1) the median time it takes for exception applications to be received by RRBs is 3 days, with significant regional variation for all requested statuses, 2) more than 25% of all patients with exception requests received heart transplants before their applications were even received by their respective RRBs, and 3) 115 people received transplants with exception requests that were ultimately denied, bypassing more than seven thousand other potential transplant recipients during the match runs.

The total number of transplants that occurred before the receipt of exception applications by the RRBs is likely an underestimate of how many transplants actually occur before final decisions are made. According to OPTN policy,^10^ the primary RRB representative initially assigned to a case has up to 3 days from receipt of an exception application to cast a vote. If the representative is unable to do so, the case is sent to an alternate representative, who then has an additional 4 days. Subsequently, all RRB representatives cast votes with final approval or denial decided by the majority. In the event of tiebreakers, the RRB chair must cast a vote, which may require additional time. Indeed, in our sensitivity analysis, by estimating the “review” date was 3 days after application receipt, transplants increased by more than 30% for patients with denials and nearly 70% for those with approvals. The fact that 115 patients with denied requests received transplants during the “travel” time is concerning because they bypassed thousands of other listed patients with spuriously elevated priority. However, the high rate of transplant among patients whose exception requests were eventually approved also raises doubts about whether RRBs can meaningfully adjudicate waitlist priority through exceptions.

The reasons for retrospective review of exception applications are not well-known. One potential motivation for this system is to avoid high waitlist mortality among patients who desperately need transplants and cannot afford to wait while their exception applications are reviewed prospectively. However, patients with status exceptions generally have lower medical urgency compared to their counterparts who meet standard listing criteria.^8,9^ In fact, our study demonstrated very low rates of death or removal for deterioration before application receipt and final decisions were rendered in our sensitivity analysis. Nonetheless, policy initiatives to reduce time to application receipt and review by RRBs may help further reduce this mortality.

As the OPTN deliberates on how to best review status exception requests, high rates of transplantation happening before applications for status exceptions are reviewed should be avoided. Centralization of exception review has already occurred in the pediatric heart allocation system with the National Heart Review Board’s implementation, which has been associated with reduced variation by center in status 1A exceptions.^12^ However, we argue that switching the system from retrospective to prospective review of exception requests is needed. Some inspiration can be taken from the liver allocation system. In May 2019, the OPTN established the National Liver Review Board, which centralized the prospective review of non-standardized Model For End-Stage Liver Disease (MELD) score exception requests and has been regularly updating comprehensive and transparent guidelines on applying for and reviewing applications.^13^ These significant changes have led not only to shorter times from application submission to decision but also significantly decreased approval rates without impacting waitlist mortality.^14^ A potential first step in switching to prospective review of applications for exceptions in the heart allocation system is to utilize the US Candidate Risk Score (US-CRS), a recently published continuous medical urgency score that is more accurate in rank-ordering candidates than the current 6-status system based on predicted 6-week waitlist mortality.^15^ US-CRS scores do not rely on hemodynamic measurements or choice of therapies and are heterogeneous among patients with status 1 and 2 exceptions^16^, suggesting wide variation in true medical urgency among exception candidates. As a result, US-CRS scores could provide an efficient method of determining which patients applying for exceptions truly have high medical urgency when applications are reviewed up front.

This study has several limitations. First, we were unable to calculate the time between application submission and actual review by the RRBs because the review date was not available in the SRTR data files. However, we demonstrated a high rate of transplantation even before the RRBs received the exception applications, and we performed a sensitivity analysis estimating a review date of 3 days after receipt. Second, our estimation of the predicted statuses without exceptions assumes that transplant centers would not have recorded additional hemodynamic measurements or made other clinical changes had they known immediately upon application submission that the exception requests were denied. Because status listings for inotropic support and temporary mechanical circulatory support (MCS) are highly dependent on the specific timing of recorded hemodynamic data, it is likely that the predicted statuses would be different if centers obtained more updated data knowing earlier about application denial from the RRBs.

### Conclusions

More than 25% of adult heart transplant candidates with submitted exception requests received heart transplants before their applications were even received by their respective RRBs, including those who have applied for exceptions that are ultimately denied. This raises significant concerns about the efficacy and fairness of retrospective review for the allocation of scarce donor hearts.

## Data Availability

All of the data files used for study in this manuscript are available upon reasonable request to the Scientific Registry of Transplant Recipients, who can be contacted via this. To obtain access to such data, one must typically submit a research proposal in order to receive a data use and access agreement. The statistical code written to perform the analyses in the manuscript are available here in this Github repository: https://github.com/danieljaechulahn/Retrospective-Heart-Exceptions

## Abbreviations

HRSA: Health Resources and Services Administration
OPTN: Organ Procurement and Transplantation Network
SRTR: Scientific Registry of Transplant Recipients
PTR: Potential Transplant Recipient
RRB: Regional Review Board

## Author Contributions

Dr. Ahn, Dr. Parker, and Dr. Sasaki had full access to the data in the study and take responsibility for the integrity of the data and the accuracy of the data analysis.

*Concept and design:* Ahn, Khush, Parker, Sasaki

*Acquisition, analysis, or interpretation of data:* Ahn, Nakayama, Attia, Eap, Parker, Sasaki

*Drafting of the manuscript:* Ahn

*Critical review of the manuscript for important intellectual content:* All authors.

*Statistical analysis:* Ahn, Parker

*Obtained funding:* Parker, Sasaki.

*Administrative, technical, or material support:* Parker, Sasaki.

*Supervision:* Khush, Parker, Sasaki

## Disclaimer

The interpretation and reporting of these data are the responsibility of the authors and in no way should be seen as an official policy of or interpretation by the SRTR or the US government.

