## Supplemental Figure 1 for "High Rate of Transplantation Prior to Review of Status Exception Requests among Adult Heart Transplant Candidates"

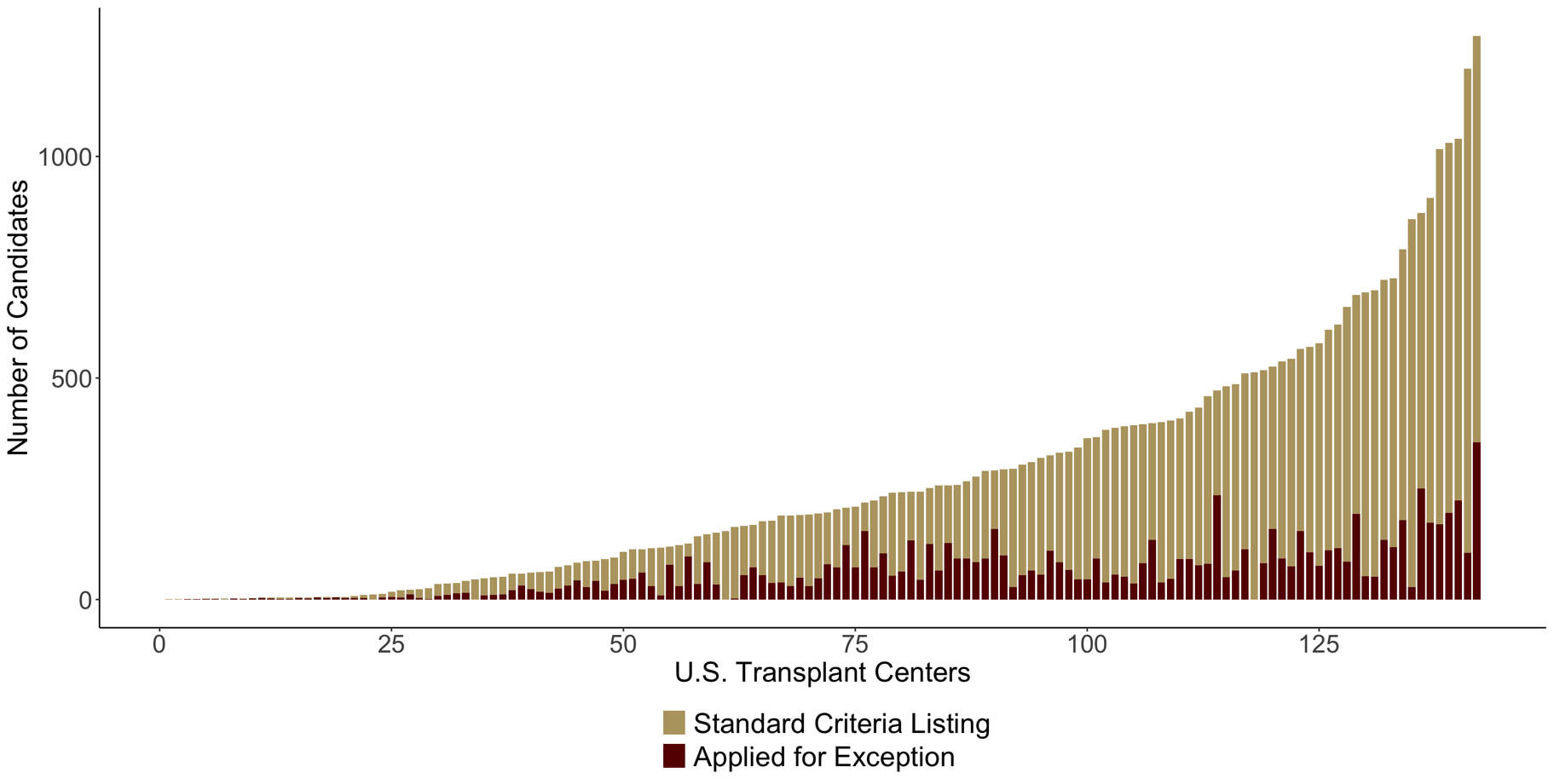
**Supplemental Figure 1**: Distribution of proportion of candidates with submitted exception applications, stratified by US transplant center.
