## Supplemental Figure 3 for "High Rate of Transplantation Prior to Review of Status Exception Requests among Adult Heart Transplant Candidates"

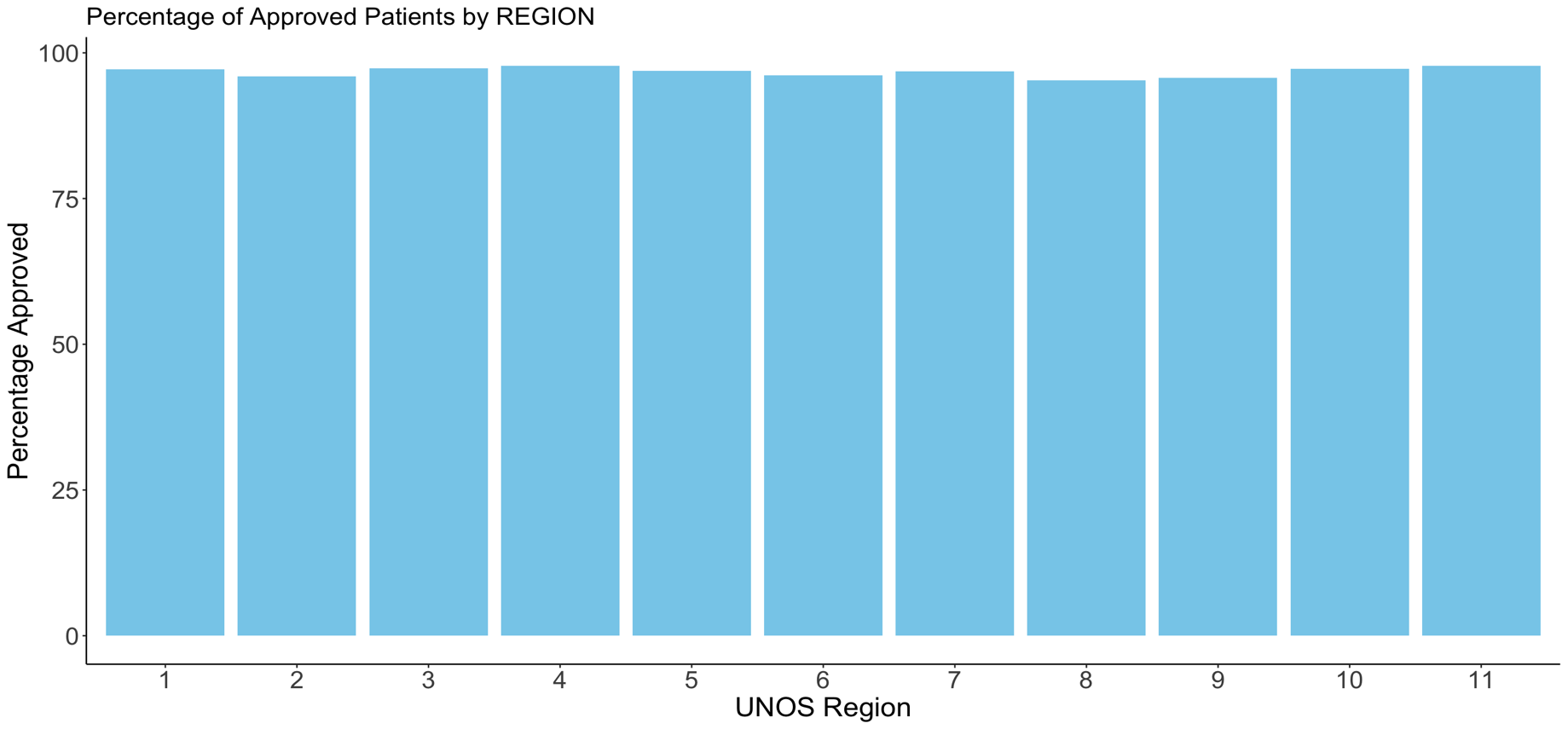
**Supplemental Figure 3**: Proportion of adult heart transplant candidates with approved exception applications, stratified by UNOS region
