## Supplemental Figure 4 for "High Rate of Transplantation Prior to Review of Status Exception Requests among Adult Heart Transplant Candidates"

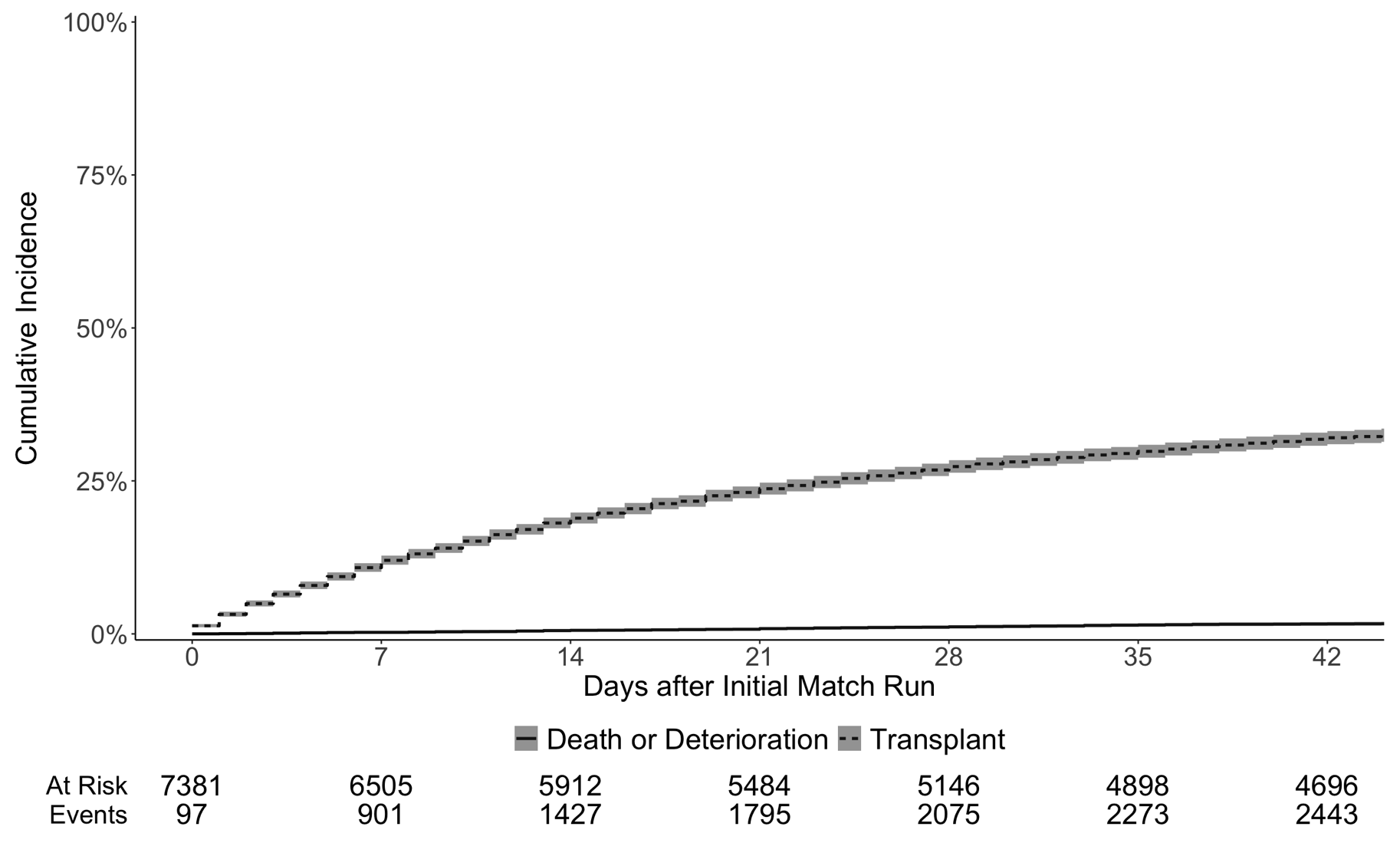
**Supplemental Figure 4**: Cumulative incidence within 6 weeks of death or removal for clinical deterioration, treating transplantation as a competing event, of potential transplant recipients bypassed by the 115 candidates who obtained heart transplants with status exceptions that were eventually denied by the regional review boards.
